# Evidence from human placenta, ER-stressed trophoblasts and transgenic mice links transthyretin proteinopathy to preeclampsia

**DOI:** 10.1101/2022.01.12.22269156

**Authors:** Shibin Cheng, Zheping Huang, Sayani Banerjee, Joel N. Buxbaum, Surendra Sharma

## Abstract

We have demonstrated that protein aggregation plays a pivotal role in the pathophysiology of preeclampsia (PE) and identified several aggregated proteins in the circulation of PE patients, most significantly the serum protein transthyretin (TTR). Here we show robust accumulation of TTR aggregates in the placentas of women with early-onset PE (e-PE). TTR aggregation was inducible in primary human trophoblasts (PHTs) and the TCL-1 trophoblast cell line by ER stress inducers or autophagy-lysosomal disruptors. Hypoxia/reoxygenation (H/R) of cultured PHTs increased intracellular BiP, phosphorylated IRE1α, PDI and Ero-1, all markers of the UPR, and the apoptosis mediator caspase-3. Blockade of IRE1α inhibited H/R-induced upregulation of Ero-1 in PHTs. Excessive UPR was observed in the PE placenta. Further, pregnant mice, overexpressing transgene encoded wild type human TTR, displayed aggregated TTR in the junctional zone of the placenta and PE-like features including hypertension, proteinuria, intrauterine growth restriction, kidney injury, and elevated levels of the PE biomarkers serum sFlt-1 and endoglin. High Resolution Ultrasound analysis revealed low blood flow in uterine and umbilical arteries compared to that found in wild type pregnant mice. On the other hand, loss of mouse TTR function did not cause any pregnancy abnormalities in *Ttr*^-/-^ mice. These observations in the PE placenta, cultured trophoblast cells and TTR transgenic mice indicate that TTR aggregation is an important causal contributor to PE pathophysiology.

## INTRODUCTION

Preeclampsia (PE) is a leading cause of maternal and fetal morbidity and mortality, affecting 3-8% of all pregnancies.^1-6^ Based on clinical features, PE is classified into early-(delivery <34 weeks, e-PE) and late-onset (delivery ≥34 weeks, l-PE) pregnancy complication.^7,8^ The clinical features include hypertension, proteinuria, glomerular endotheliosis, and fetal growth restriction.^1-9^ PE may be life-threatening for the mother with multi-organ complications including renal, cardiac and neurologic involvement.^2,6^ It is not known what triggers the onset of PE. Placenta-specific insufficiency and defective trophoblast invasion of the endometrium resulting in poor spiral artery remodeling have been demonstrated to be contributing factors.^1-9^ Although normal pregnancy-associated mild, physiological endoplasmic reticulum (ER) stress and low oxygen tension are associated with the local endovascular milieu required for trophoblast growth and migration,^1,2,9,10^ it is possible that in PE patients, chronic or increased local hypoxia/ischemia, excessive UPR activation and acute or chronic effects on cellular machinery create a degree of stress that exceeds the proteostatic capacity of placental trophoblasts causing placental insufficiency.

We and others have recently demonstrated that PE is associated with placental proteinopathy (a pathological condition caused by misfolded and aggregated proteins), a hallmark feature of neurodegenerative diseases such as Alzheimer’s disease and the systemic amyloidoses.^2,11-14^ We have also demonstrated that impaired autophagy coupled with poor lysosomal biogenesis and pyroptosis are critical pathologies in the PE placenta.^16-18^ These observations raise the question of whether excessive placental ER stress and/or hypoxia-oxygenation (H/R) induce PE-associated pathophysiological alterations, such as protein aggregation and an increase in circulating anti-angiogenic soluble factors (soluble fms-like tyrosine kinase-1 (sFlt-1) and soluble endoglin (sEng)), inflammatory cytokines, exosomes and alarmins (e.g. cell free fetal DNA, HMGB1, and uric acid).^19-24^

We have previously established a serum-based “humanized” mouse model of PE that recapitulates all the features associated with human PE.^25^ The model involves the administration of sera from women with severe PE to pregnant wild-type or IL-10^-/-^ mice. Comparative proteomic screening of normal pregnancy and PE sera revealed dysregulated transthyretin (TTR) as a potential pathogenic factor in PE.^2,11^ Our studies have revealed that aggregated TTR is a key cargo of nano-vesicles released from PE placental explants, suggesting that toxic protein aggregates can be released from the PE placenta into the maternal circulation and may act as alarmins, leading to systemic inflammation and endothelial dysfunction.^12^ Indeed, using a novel protein aggregation detection assay, we have found significantly higher levels of TTR aggregates in sera from women with PE than those obtained from women undergoing normal pregnancy.^26^ Based on our observations that autophagy is impaired in the PE placenta, we utilized an autophagy-deficient trophoblast line to establish this novel assay of protein aggregate detection. However, it is not clear what leads to impaired autophagy and whether aggregated proteins, particularly TTR, contribute to the pathogenesis of PE.

Monomer TTR is a 127 amino acid protein encoded by a single gene on human chromosome 18.^27^ Functional TTR is a 55-kDa homo-tetramer that serves as a minor transporter of thyroxine and the major carrier of retinol binding protein charged with retinol in the plasma. Although mainly synthesized in the liver and the choroid plexus of the brain, TTR synthesis has also been noted in the pancreas, in neurons under stress, and skin.^28-30^ TTR is also expressed in the trophoblast layer of the human placenta as early as 6 weeks of pregnancy and throughout gestation.^28-30^

TTR plays an important role in embryological development of the brain through transplacental delivery of thyroxine. Thyroxine is crucial to embryological development of the fetal central nervous system. The fetus relies solely on maternal thyroxine before it begins to secrete thyroid hormones at around 16 weeks gestation.^30^ In addition, TTR has been shown to regulate angiogenesis in endothelial cells^31^, promote nerve regeneration after injury^32^, and prevent the progression of Alzheimer’s disease by inhibiting amyloid-β peptide (Aβ) fibril formation and by sequestering/clearing Aβ aggregates.^33,34^ Native TTR normally circulates as a moderately stable tetramer with a low rate of dissociation into its monomers (15 kD). The free monomers are aggregation-prone, particularly if they carry a mutation or have undergone post-translational oxidative change.^35^ TTR oligomers are cytotoxic to cells in tissue culture.^35^ TTR-derived fibril deposition has been associated with three systemic amyloid diseases, two associated with variant protein structures, i.e. familial amyloidotic polyneuropathy, familial amyloidotic cardiomyopathy, and one in which the protein structure is wild type, senile systemic amyloidosis.^35^

Intracellular misfolded and aggregated proteins are removed by the unfolded protein response (UPR) machinery and the autophagy-lysosomal degradation system respectively.^2,16,17^ It is possible that accumulation of aggregated proteins results from failed UPR and protein degradation mechanisms or that the generation of misfolded protein quantitatively exceeds the intrinsic capacity of the proteostasis network in the trophoblast. We hypothesize that persistent environmental stress factors, such as chronic low oxygen tension, lead to increased intracellular protein misfolding resulting in excessive ER stress, which exhausts the capacity of UPR and impairs autophagy-lysosomal degradation machinery, resulting in the accumulation of misfolded and aggregated TTR in the PE placenta. Alternatively, it is possible that the misfolding, aggregation and deposition of TTR and other molecules in the placental environment compromise placental blood flow.

Here we focused on the placenta from patients with severe e-PE which show impairment of the UPR and autophagy-lysosomal pathways. In a cellular model of PE pathophysiology induced by H/R, we found increased TTR aggregation. Further, when we exposed cultured trophoblast cells to ER stress inducers or an autophagy-lysosome disruptor, we also observed increased cellular TTR aggregation. We found similar changes in the placentas of pregnant mice genetically programmed to over-express wild type human TTR, further supporting the notion of a significant contributory role of aggregated TTR in PE pathophysiology.

## METHODS

The authors declare that all supporting data are available within the article and its Supplemental Material.

### Human subjects

PE was diagnosed as new-onset systolic and diastolic blood pressures of >140 and 90 mm Hg, respectively, and proteinuria (>300 mg of protein in a 24-hour urine collection or a random urine protein/creatinine ratio of >0.3) after 20 weeks’ gestation. Severe PE was further defined by systolic and diastolic blood pressures of >160 and 110 mm Hg, respectively. Eight women with severe e-PE (<34 gestational weeks) PE and 8 women with gestational age-matched pregnancy were enrolled under protocols approved by the Institutional Review Boards of Women and Infants Hospital of Rhode Island (Providence). Patient demographic information included for the study are shown in Supplemental Table S1. For placental sample collection, a 1cm^3^ specimen was removed from the middle of the placenta and vigorously rinsed in chilled phosphate-buffered saline (PBS) solution. After removing any additional blood from placental tissue, a portion was stored at -80° C until further use; the remaining tissue was fixed in 10% formalin for immunohistochemical analysis. Informed consent was obtained from all subjects.

### Antibodies and reagents

Commercial antibodies included: rabbit anti-human transthyretin antibody (A0002, Agilent Dako, Santa Clara, CA), rabbit anti-BiP (Abcam, Cambridge, MA), rabbit anti-pIRE1a (Abcam, Cambridge, MA), rabbit anti-Ero-1 (Cell Signaling Technology, MA), rabbit anti-PDI (Abcam, Cambridge, MA), rabbit anit-LAMP 1 (Abcam, Cambridge, MA), rabbit anti-albumin (Abcam, Cambridge, MA) rabbit anti-ubiquitin (Santa Cruz, CA), rabbit anti-caspase-3 (Cell Signaling Technology, MA), rabbit anti-SQSMT1/p62 (MBL), mouse anti-β-actin (Cell Signaling Technology, MA), goat anti-rabbit or mouse HRP-conjugated IgG (Cell Signaling Technology, MA), donkey anti-rabbit Alexa 488 or 594 (ThermoFisher Scientific, MA). STF 083030 (an IRE1a inhibitor) was purchased from, Santa Cruz, CAS (307543-71-1)

### Cell lines and culture

Human primary trophoblasts (PHTs, purchased from ScienCell Research Laboratories, Carlsbad, CA) were cultured in TM medium (ScienCell Research Laboratories, Carlsbad, CA) supplemented with 10% FBS, growth factors (ScienCell Research Laboratories, Carlsbad, CA), 100 U/ml penicillin and 100 µg/ml streptomycin (GIBCO, 15140, ThermoFisher Scientific, MA) at 37°C in a 5% CO_2_ atmosphere. Third trimester trophoblast cells (TCL-1) were grown at 37 °C in media supplemented with 10% fetal bovine serum and 100 U/ml penicillin and 100 µg/ml streptomycin (GIBCO, 15140, ThermoFisher Scientific, MA).

### Hypoxia-reoxygenation treatment

PHTs and TCL-1 cells were washed with serum-free media and then exposed for 72 h to normal or low oxygen tension (<1% O_2_) using a hermetically enclosed incubator (Thermo Electron, Marietta, OH, USA) with continuous digital recording of atmospheric oxygen using a sensor connected to a data acquisition module (Scope; Data Translation, Marlboro, MA, USA). The media were pre-equilibrated to the gas mixture before addition to the culture plate. At day 3, hypoxia-treated cells were incubated for 2-3 h under normoxic condition for reoxygenation treatment. The cells were fixed with 4% paraformaldehyde or lysed in RIPA buffer containing 25 mM Tris-HCl, 150 mM NaCl, 1% sodium deoxycholate, 1% NP40, 0.1% SDS, protease inhibitor cocktail (0469316001, Roche, Switzerland), and 1% phosphatase inhibitor (P5726, Sigma-Aldrich, St. Louis, MO).

### Immunofluorescence

Cells were fixed for 10-15 min with 4% paraformaldehyde and permeabilized in blocking buffer containing 0.1% Triton X-100, 3% BSA, and 3% normal goat or donkey serum in PBS (pH 7.4) for 25 min at room temperature. Cells were then incubated overnight with primary antibodies diluted in blocking buffer. After several washes in PBS, cells were incubated at room temperature with donkey anti-rabbit Alexa 488 or 594 for 1 h. The sections or cells on the coverslips were mounted with anti-quenching mounting medium with DAPI (Vector Laboratories Inc., Burlingame, CA) and observed using a Nikon Eclipse 80i fluorescence microscope (Tokyo, Japan) or a confocal microscope (Nikon, A1R, Japan) with a Texas Red filter set for Alexa 594 antibodies and a FITC filter set for Alexa 488 antibodies, respectively. Images from both control and treated cells were acquired using identical settings. The specificity of staining was tested by omitting the primary antibody or using normal rabbit IgG or TTR protein-neutralized primary antibody. Images were processed with brightness/contrast adjustment. Linear adjustment of contrast and brightness was applied to all entire images equally using Photoshop CS6 (Adobe Inc., San Jose, CA).

### Detection of TTR aggregates

Paraffin-embedded sections of placental tissues from term-matched normal pregnant and preeclamptic women were de-paraffinized and stained with ProteoStat dye for 5 min at room temperature according to a modified previously described protocol.^16^ After several washes, sections were immunostained overnight at 4 °C for TTR with rabbit anti-TTR antibody (A0002, Agilent Dako, Santa Clara, CA) and then incubated for 1 h with goat anti-rabbit Alexa 488 at room temperature. Nuclei were stained with DAPI. For cell lines, TCL-1 cells were plated onto glass coverslips, treated with various agents or hypoxia/normoxia, then fixed with 4% paraformaldehyde at various time points. Cells were permeabilized with 0.5% Triton X-100, processed for staining with ProteoStat dye (Enzo Life Sciences, NY) according to manufacturer’s instruction and then counterstained for TTR with rabbit anti-human TTR antibody (A0002, Agilent Dako, Santa Clara, CA). TTR immunoreactivity was visualized with goat anti-rabbit Alexa 488 secondary antibody (ThermoFisher Scientific, MA). The sections or cells on the coverslips were mounted with anti-quenching mounting medium with DAPI (Vector Laboratories Inc., Burlingame, CA) and observed using a Nikon Eclipse 80i fluorescence microscope (Tokyo, Japan) or a confocal microscope (Nikon, A1R, Japan) with a Texas Red filter set for the ProteoStat dye (the excitation/emission wavelengths: 530/560-700 nm) and a FITC filter set for Alexa 488 antibodies, respectively.

### Immunoblotting

Cells were lysed in RIPA buffer containing 50 mM Tris, 150 mM NaCl, 0.5% sodium deoxycholate, 1% Triton X-100, 1mM PMSF and protease inhibitor cocktail (Sigma-Aldrich, St. Louis, MO). Protein concentration was measured with the BCA method. The protein extract was electrophoresed in a 4-15% gradient gel under non-reducing conditions (without SDS and 2-ME in the sample buffer and no boiling). Under reducing conditions, the protein extract was mixed with denaturing sample buffer (5% SDS, 2.5% 2-ME and 100 mM DTT), boiled and separated using a 4-15% SDS-PAGE. The transferred PVDF membrane was blocked for 1 h with blocking solution containing nonfat dried milk (5%) prepared in phosphate-buffered saline containing 0.05% Tween 20. Proteins were detected by probing overnight with primary antibodies diluted in the blocking solution, secondary goat anti-rabbit horseradish peroxidase for 1 h at room temperature, and finally visualized by incubating with enhanced chemiluminescence substrate (ThermoFisher Scientific, MA).

### In vivo studies

Transgenic mice over-expressing human TTR (huTTR mice) and mice lacking endogenous *Ttr* gene (TTR KO) were generated as described previously.^37^ The transgene construct contains 90-100 copies of the normal human TTR gene and all the known regulatory elements required for tissue specific expression. The plasma of the mice contains 1-3mg TTR/dl.^37-39^ All procedures were performed according to protocols approved by the Institutional Animal Care and Use Committee (IACUC) at The Scripps Research Institute. IL-10^−/−^ mice with a C57BL/6 background were purchased from The Jackson Laboratory (Main, USA). All animal protocols were approved by the Lifespan Institutional Animal Care and Use Committee. All mice were allowed free access to food, water, and activity. All mice were used between 7 and 9 weeks of age at the time of pregnancy. The day of vaginal plug appearance was designated gestational day (gd) 0. On gd 10, native TTR or TTR aggregates were i.p. injected at a 40 mg/kg dose in IL-10^-/-^ mice. On gd 16, pregnant IL-10^-/-^, huTTR mice, TTR KO and wild type mice were transferred to metabolic cages for 24-hour urine collection for assessing proteinuria. Total urinary albumin was measured using Albumin (mouse) ELISA kit (ALPCO Diagnostics, Germany). To normalize the albumin, urinary creatinine was measured using Metra Creatinine Kit (Quidel Corporation, San Diego, CA), according to the manufacturer’s protocol. Proteinuria was represented as the ratio of urinary albumin/creatinine. On gd17, blood pressure was recorded by the tail-cuff method as described earlier.^9,11^ The animals were then euthanized, and fetal weights were recoded. The kidneys and placentas were fixed in 10% formalin and paraffin-embedded for section. Sections from the kidney and placenta were stained with H&E and PAS for histopathological examination as previously described.^9,11^

### ELISA of serum TTR, sFlt-1 and sEng concentrations

Sera were obtained from human wild type TTR transgenic and wild-type mice on gd 17 at age of 7-9 weeks. Diluted sera (1:20,000 times for TTR assay, 1:10 for sFlt-1 detection and for sEng) were assessed using TTR ELISA kit (Immunodiagnostic AG, Germany) or mouse VEGF R1/Flt-1 or sEng kit (R&D Systems, Minneapolis, USA) as described in manufacturers’ instructions. The optical density of colorimetric reaction substrates of the samples was read using iMark™ Microplate Absorbance Reader (Bio-Rad Laboratories, CA).

### Doppler ultrasound analysis

On gd 16, uterine and umbilical artery blood flow were assessed using Doppler ultrasonography (Vevo3100, VisualSonics, Canada). Peak systolic and end-diastolic velocities were measured from at least 3 consecutive cardiac cycles that were not affected by motion caused by maternal.^36^ The peak systolic and diastolic velocities of uterine and umbilical arteries were calculated to evaluate alterations in blood flow using the software (Vevo LAB 3.1.1, VisualSonics, Canada).

### Statistical analysis

Statistical comparisons were conducted using Students *t*-test. The results were expressed as the mean ± SEM. P values of less than 0.05 were considered significant.

## RESULTS

### Deposition of TTR aggregates in the placenta from e-PE deliveries

To provide direct evidence for aggregated TTR deposition in the placenta from PE patients, we employed dual staining using TTR antibody in combination with ProteoStat dye, a rotor dye that uniquely binds aggregated proteins.^16,26^ We have recently reported a novel blood test for PE using the unique ability of ProteoStat to bind to aggregated proteins.^26^ This assay detects TTR, amyloid β-42, phosphorylated tau, and α-synuclein aggregates in sera from patients with PE and Alzheimer’s disease.^26^ We show here that ProteoStat also works well with tissue and cultured cells. Placental tissue sections from e-PE and gestational age-matched deliveries were stained with ProteoStat dye and then immunostained for TTR. As shown in Figure 1, the PE placenta displayed much more intense staining for TTR and ProteoStat fluorescence relative to control placenta. The staining was primarily localized to the trophoblast layer of microvilli and the extravillous trophoblasts (EVT). Notably, the TTR immunofluorescent signal co-localized with the ProteoStat signal, indicating the aggregated nature of TTR in the PE placenta. It was recently reported that albumin, a plasma protein abundant in circulation and placenta, was detected in aggregated protein complexes in urine samples of PE patients.^15^ We examined whether aggregated albumin could be identified in the placental protein aggregates using the ProteoStat approach. Our dual staining analysis shows strong positive albumin staining in the trophoblast layer and in blood vessels in the placenta from both control and preeclampsia deliveries. However, while the e-PE placenta exhibited higher levels of ProteoStat fluorescence, albumin was not detected in the aggregated form as evidenced by a lack of co-localization of albumin with the ProteoStat signal (Figure S1 in the Supplemental Material). It is possible that aggregated albumin in the urine is related to PE-associated renal dysfunction.

**Figure 1.**
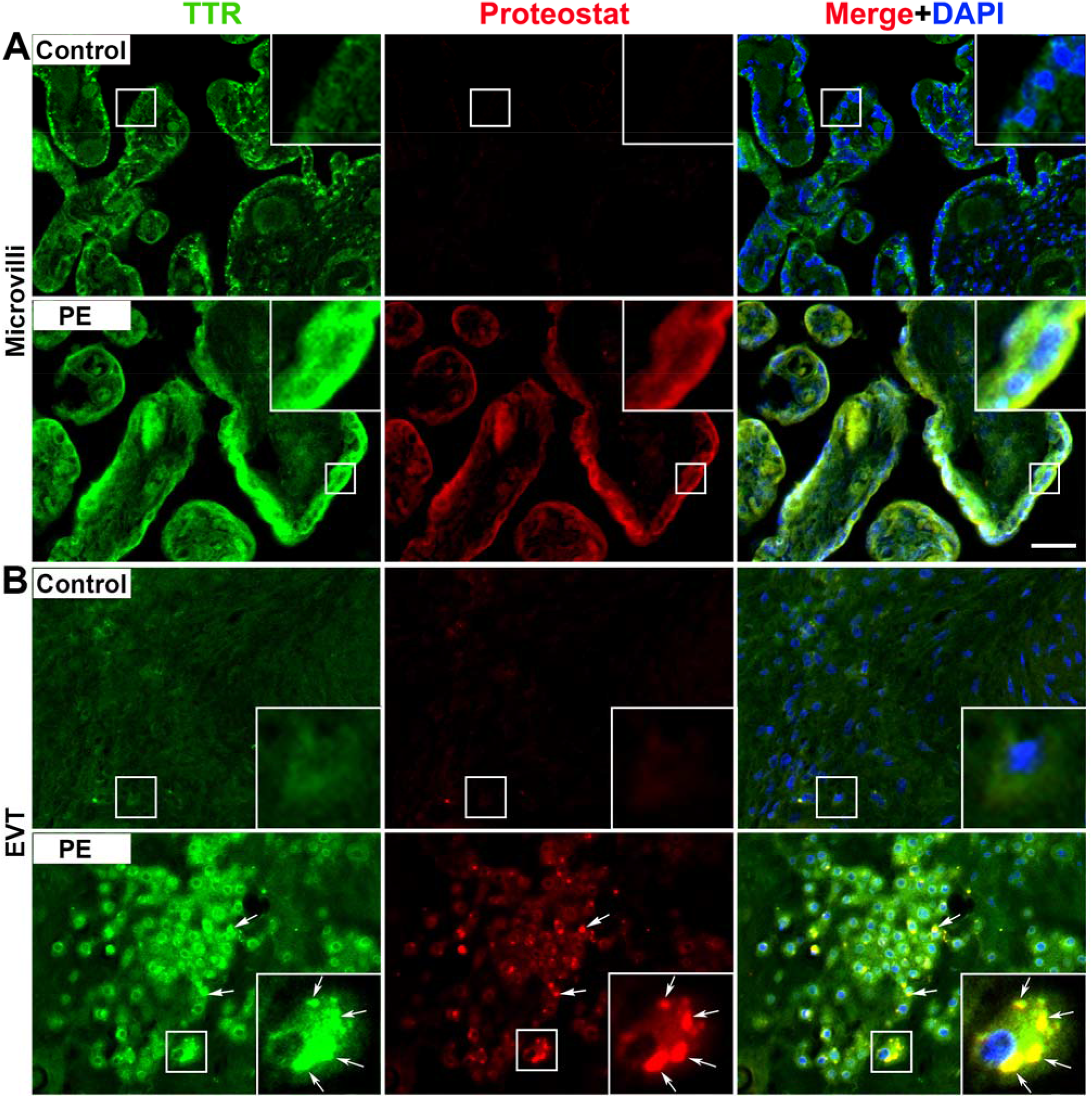
Colocalization of TTR immunoreactivity with ProteoStat signal in the placenta from e-PE and gestational age-matched control. Placenta tissue sections were immunostained for TTR (green) and co-stained with ProteoStat dye (red). The nuclei were stained with DAPI (blue). Yellow color showed colocalization of TTR green signal and ProteoStat red signal after red, green and blue channels were merged. **A**, TTR aggregates in the trophoblast layer of microvilli. **B**, TTR aggregates in the extravillous trophoblasts (EVT). Inserts were magnified images of boxed areas. The images were representatives of at least three independent experiments. Arrows indicated puncta-like TTR aggregates. Scale bar: 50 µm

### MG132 (proteasome inhibitor), Brefeldin A (ER stress inducer), and Chloroquine (lysosomal inhibitor) induce accumulation of TTR aggregates in primary human trophoblasts and TCL-1 human trophoblast cell line

Degradation of misfolded and aggregated proteins through the UPR and autophagy-lysosomal pathways is a crucial adaptive response under ER stress.^2,16,17^ To delineate the mechanism(s) underlying the accumulation of TTR aggregates in the PE placenta, we tested whether TTR aggregates accumulated in human trophoblasts when proteasomal and autophagy-lysosomal pathways were disrupted or trophoblasts were subjected to excessive ER stress. We treated primary human trophoblasts (PHTs) and TCL-1 cells (third trimester extravillous trophoblast cell line) overnight with vehicle or MG132, a proteasome inhibitor, chloroquine, an inhibitor of lysosome acidification or brefeldin A (BFA), which inhibits vesicle transport from the ER to the Golgi, and examined the state of TTR in the treated cells using immunofluorescence in combination with ProteoStat staining. As depicted in Figure S2 (in Supplemental Material), strong TTR immunoreactivity and robust ProteoStat fluorescence were observed in MG132, chloroquine, or BFA-treated PHTs compared to untreated control cells. Importantly, the TTR signal co-localized with the ProteoStat fluorescence in treated cells, indicative of the aggregated nature of TTR. Similarly, TTR aggregate accumulation was observed in TCL-1 cells treated with MG132, BFA or chloroquine (Figure S3 in the Supplemental Material). These results demonstrated that TTR aggregated in PHTs when cellular proteasomal and autophagy-lysosomal degradation machineries were disrupted or when trophoblasts were subjected to excessive ER stress.

### Hypoxia-reoxygenation (H/R) exposure induces TTR aggregation in PHTs and TCL-1 cells

Our prior studies revealed that chronic low oxygen tension induced excessive ER stress, which, in turn, impaired the autophagy-lysosome machinery in trophoblasts.^16^ Thus, we next employed a cellular model using H/R to determine whether exposure to H/R would lead to accumulation of TTR aggregates in trophoblasts. PHTs or TCL-1 cells were cultured in serum-free media under H/R in a calibrated hypoxia chamber to maintain 1% O_2_ and then lysed for western blotting or fixed for immunofluorescent staining at day 1, 2 or 3 as described in Methods. Similar experiments were performed with PHTs cultured under normoxic conditions. Protein extracts were separated using western blotting under non-reducing conditions. Immunoblotting and quantitative analysis demonstrated enhanced levels of TTR aggregates with high molecular weight protein moieties in TCL-1 cells at days 2 and 3 (Figure S4A and B in the Supplemental Material). In contrast, no high molecular weight albumin aggregates were detected in H/R-treated TCL-1 cells (Figure S4C in the Supplemental Material). Based on these results, we examined TTR aggregation in normoxia-and H/R-treated PHTs at day 3 by western blotting and immunohistochemistry. Significant TTR aggregation, as depicted by high molecular weight protein bands stained with the anti-TTR antibody, was detected in H/R-exposed PHTs (Figure 2A). Quantification of the signal showed that H/R significantly increased the abundance of TTR aggregates and oligomers in PHTs (Figure 2B). Co-staining with anti-TTR antibody and ProteoStat dye revealed higher levels of TTR immunoreactivity and ProteoStat fluorescence in H/R-treated PHTs (Figure 2C) and TCL-1 cells (Figure S4D in the Supplemental Material) compared to controls. Co-localization of TTR with the ProteoStat dye was observed only in H/R-treated cells. ProteoStat dye staining was not detected in normoxia-treated cells (Figure 2C and Figure S4D in the Supplemental Material). These results indicate that the H/R induced accumulation of TTR aggregates in PHTs and TCL-1 cells.

**Figure 2.**
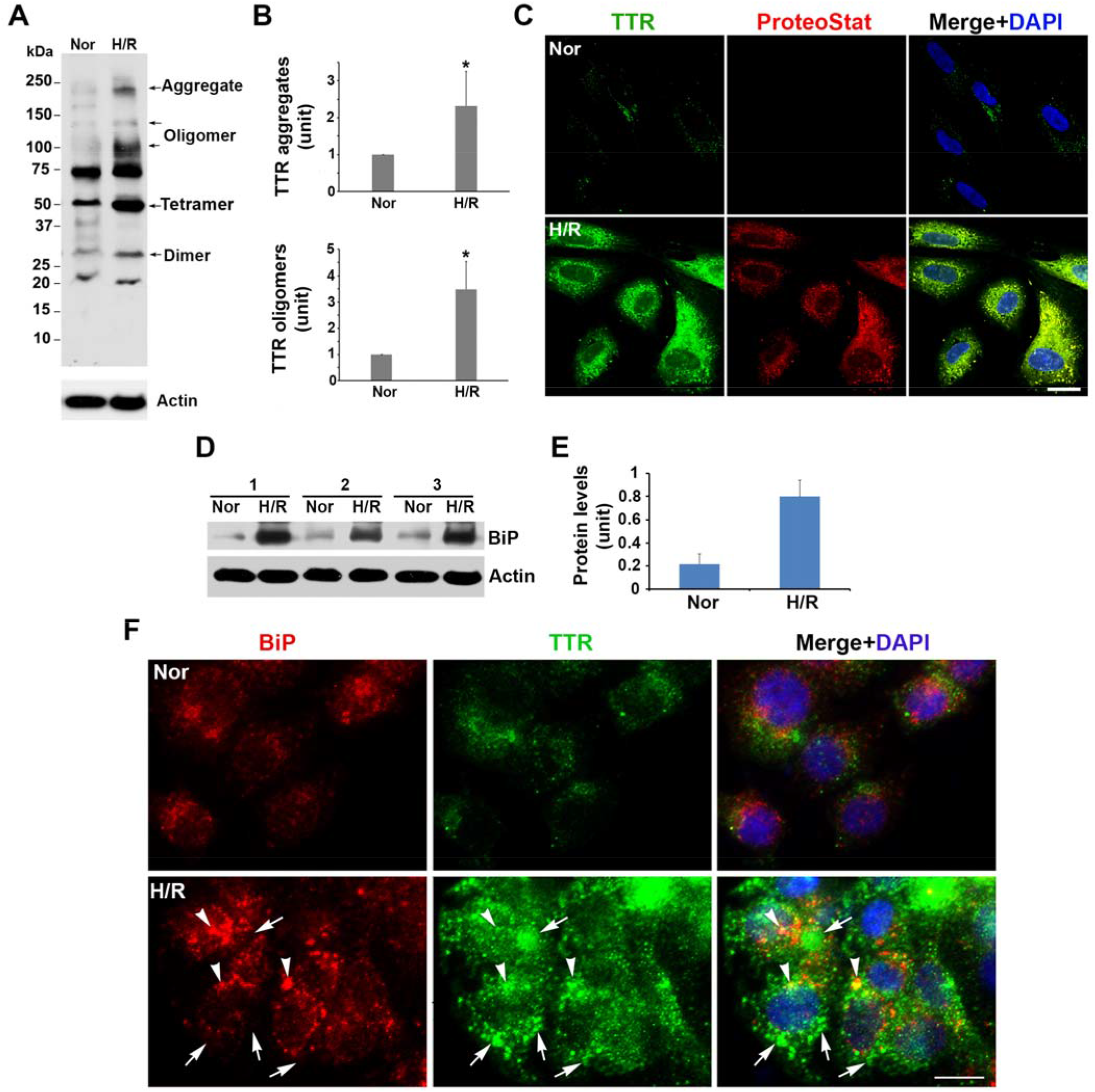
H/R exposure induces TTR aggregation, increases the abundance of BiP and decreases its colocalization with TTR immunoreactivity in PHTs. The cells were treated with normoxia or H/R and lysed or fixed at 3 days. Protein extracts were separated using western blotting, and fixed cells were immunostained for TTR (green) combined with ProteoStat staining (red). **A, B**, Western blotting and quantitative data showed higher levels of TTR aggregates and oligomers in H/R-vs. normoxia-treated cells. Actin was used as a control of sample loading. **C**, Colocalization of TTR immunoreactivity with ProteoStat fluorescence signal in cells treated with H/R not normoxia. The nuclei were stained with DAPI (blue). The images were representatives of at least 3 independent experiments. **D, E**, western blotting and quantitative data showed a higher level of BiP in H/R-exposed trophoblasts. **F**, Representative images showed the colocalization of TTR and BiP in normoxia-treated (Nor) and H/R-treated cells (n=3). The nuclei were stained with DAPI (blue). Data were expressed as mean ± SEM and statistically analyzed by the Student’s *t-*test (n = 3). *: *p* < 0.05. Scale bar: 20 µm

### H/R treatment increases abundance of BiP but inhibits its co-localization with TTR aggregates in PHTs

We hypothesized that the accumulation of TTR aggregates might result from dysregulated UPR in trophoblasts in response to excessive hypoxic stress. Binding immunoglobulin protein (BiP), an ER lumen chaperone protein regulating the UPR pathway, is widely used as a marker for ER stress and UPR activation because binding of BiP by misfolded proteins activates the UPR.^2^ To determine the effect of chronic H/R exposure on UPR activity, we first examined the expression of BiP in PHTs cultured under the H/R conditions. As shown in Figure 2D and E, H/R-treated cells exhibited significantly higher levels of BiP compared to controls, suggesting that the UPR was activated. Under physiological UPR activation, BiP binds to misfolded proteins and guides them for proper refolding. To assess whether the activation of the UPR is excessive under the H/R conditions, we performed dual immunostaining to evaluate co-localization of BiP with TTR proteins. Our results revealed more BiP immunoreactivity and puncta-like TTR signals in H/R-exposed PHT cells than in control cells (Figure 2F). Immunostained TTR was distributed throughout the cytoplasm of H/R-treated cells. Notably, despite large amounts of BiP fluorescence signals, only a few BiP immunoreactive signals overlapped with TTR immunoreactivity and the majority of TTR proteins did not co-localize with the BiP signal in H/R-exposed PHTs, indicating that much of TTR is in the cytoplasm not in the ER lumen.

### H/R treatment enhances the contents of pIRE1α, PDI, Ero-1 and caspase-3 in PHTs

We next assessed the expression of multiple key index molecules that determine the extent of ER stress and the UPR activity in H/R-treated cells. To this end, the content of pIRE1α, PDI, Ero-1 (a component of the PERK cell death pathway of the UPR), and cleaved caspase-3 (the executioner of apoptosis) was examined in PHTs exposed to H/R. As shown in Figure 3A-E, pIRE1α, PDI, Ero-1 and cleaved caspase-3 proteins were significantly increased in H/R-exposed cells relative to control cells (*p* < 0.05), indicating that all the elements of the UPR were highly activated during exposure to H/R.

**Figure 3.**
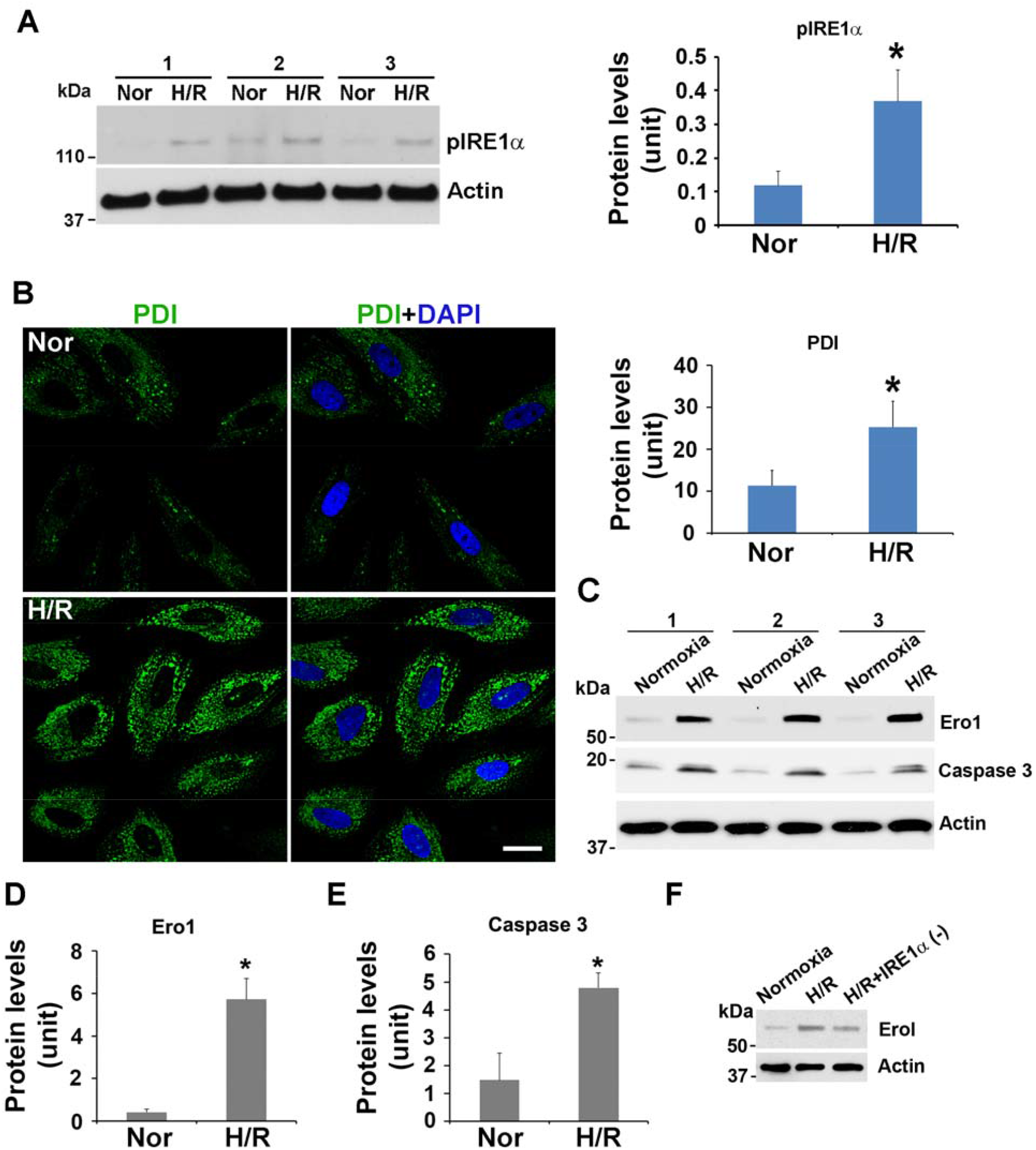
H/R treatment hyper-activates the UPR machinery in PHTs. The cells were exposed to normoxia or H/R and lysed or fixed at 3 days. **A**, Western blotting and quantitative analysis showed a higher level of pIRE1a in the cells treated with H/R vs. normoxia. **B**, Representative images using immunofluorescence staining demonstrated a higher pixel intensity of PDI (green) in H/R treated cells compared to normoxic controls (n = 3). The nuclei were stained with DAPI (blue). Scale bar: 20 µm. **C**-**E**, Western blotting and quantitative analysis showed higher levels of Ero1 and caspase-3 in H/R-treated cell relative to normoxic controls. Data were expressed as mean ± SEM and statistically analyzed by the Student’s *t-*test (n = 3). *: *p* < 0.05. **F**, Inhibitor of IRE1α attenuated the abundance of Ero1 in H/R-treated trophoblasts.

Next, we assessed whether inhibition of prolonged pIRE1α activation can attenuate the production of Ero1 in H/R-treated PHTs. Cells were treated with STF 083030, an inhibitor of IRE1α at day 2 during 3-day exposure to H/R or normoxia. As expected, inhibition of IRE1α, significantly reduced Ero-1 presence in the H/R-exposed cells compared to control cells (Figure 3F).

### H/R exposure inhibits colocalization of TTR aggregates with the lysosomal marker LAMP1 in PHTs

Our results demonstrated that disruption of the lysosome machinery by chloroquine elicited the accumulation of TTR aggregates in PHTs. We also found robust accumulation of TTR puncta-like signals in the cytoplasm of H/R-treated trophoblasts. To examine the state of the lysosomal machinery in H/R-treated PHTs, we examined the expression of lysosomal-associated membrane protein 1 (LAMP1) and colocalization of LAMP1 with TTR signal in PHTs exposed to H/R. As shown in Figure S5A (in Supplemental Material), total LAMP1 content was significantly reduced while the TTR signal is high. Further, there was little evidence for co-localization of LAMP1 with H/R-induced TTR (Figure S5B in Supplemental Material). This suggests that chronic hypoxia may impair lysosomal integrity and attenuate degradation of TTR aggregates in the lysosomes of PHTs.

### Increased abundance of BiP, pIRE1α, PDI, Ero-1, caspase-3, SQSTM1/p62 and total ubiquitinated proteins in the PE placenta

We then assessed whether the molecular events that were observed in the cellular model mimicking the PE pathophysiology induced by H/R were also seen in placental tissue from women with e-PE. To address this issue, the quantities of BiP, pIRE1α, PDI, Ero-1 and caspase-3 were evaluated in placental tissue sections from e-PE deliveries. Immunofluorescent and quantitative analyses revealed higher levels of BiP, pIRE1a and PDI in the trophoblast layer of the e-PE placenta relative to the control tissue (Figure 4A-C). Immunoblotting analysis also revealed greater abundance of Ero-1 and caspase-3 in the placenta from e-PE deliveries (Figure 4D).

**Figure 4.**
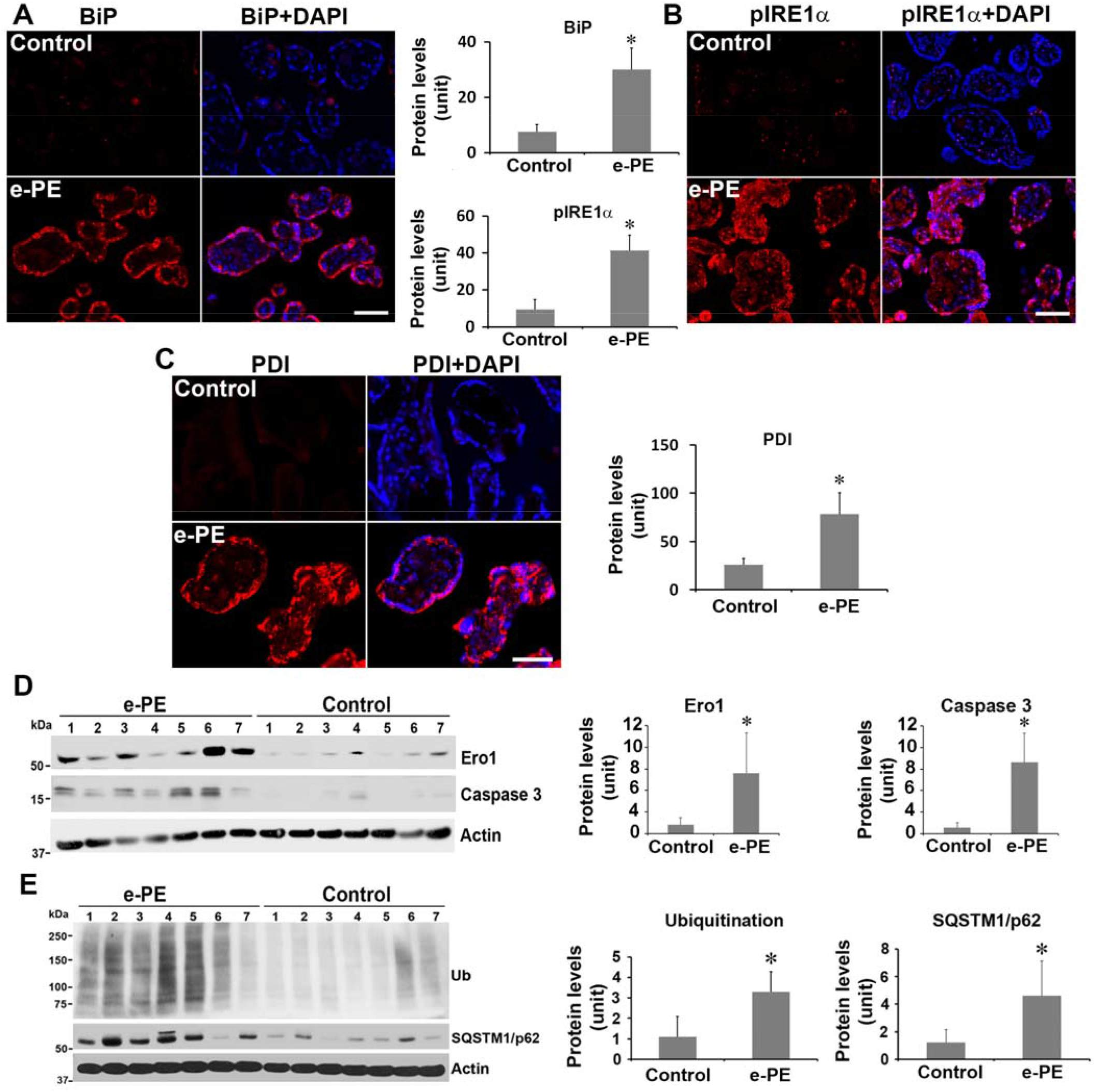
The expression of BiP, pIRE1a, PDI, Ero1, caspase-3, SQSTM1/p62 and total ubiquitinated proteins in the placenta from e-PE patients and gestational age-matched controls. **A-C**, Immunofluorescence staining and quantitative analyses for BiP (**A**), pIRE1a (**B**) and PDI (**C**) in control and e-PE placenta. The nuclei were stained with DAPI (blue). The images were representatives of 7 independent experiments. **D, E**, Immunoblotting for Ero1, caspase 3, SQSTM1/p62 and total ubiquitinated proteins, and quantitative analysis of the band intensity of these two proteins in control and e-PE placenta. Data were expressed as mean ± SEM and statistically analyzed by the Student’s *t-*test (n = 7). *: *p* < 0.05. Scale bar: 20 µm

We then investigated whether placental autophagy is impaired by examining the abundance of SQSTM1/p62, the link protein between ubiquitin-conjugated proteins and autophagy, and total ubiquitinated proteins in the placenta from e-PE deliveries. As shown in Figure 4E, the abundance of SQSTM1/p62 and total ubiquitinated proteins were strikingly higher in the PE placenta compared to control tissue. The data suggest that autophagy and ubiquitination-mediated protein degradation pathways may be compromised in the PE placenta, which may explain why TTR aggregates accumulated and were detected in sera of PE patients. These results are consistent with our previous data showing impairment of autophagy and lysosome biogenesis in the PE placenta.^16^

### Overexpression of human TTR in transgenic mice (huTTR) induces PE-like features and increases concentration of serum sFlt-1 and sEng in pregnant mice

Using a strain of transgenic mice which contains 90-100 copies of the wild type human *TTR* gene with all its known regulatory elements and shows very high tissue expression^37^, we investigated whether TTR undergoes aggregation and plays a causal role in PE pathophysiology. First, we examined the serum concentration of TTR and deposition of TTR aggregates in the placenta in huTTR mice. As previously shown, ELISA and western blotting using an antibody specific to human TTR revealed a remarkably high level of human TTR (113 mg/dl on average) in sera from huTTR mice at gd17. In contrast, no human TTR was detected in sera from wild-type (non-transgenic) mice (Figure 5A). Dual staining showed large amounts of TTR immunoreactivity and ProteoStat fluorescence in the placenta of the huTTR compared to wild-type mice, with the signal intensity significantly higher in the junctional zone. Importantly, the TTR immunoreactive signal co-localized with the ProteoStat fluorescence signal, suggesting that TTR was present in the aggregated form in the placenta of huTTR mice (Figure 5B). To determine whether TTR aggregates in the placenta had any pathophysiological effects, we evaluated huTTR mice and wild-type pregnant mice for PE-like features. As shown in Figure 5C, pregnant huTTR mice had higher systolic blood pressure and ratio of albumin to creatinine relative to wild-type controls and their pups had lower average fetal weights. In addition, glomerular endotheliosis was observed in the kidney tissue of pregnant huTTR mice but not wild-type mice (Figure 5D).

**Figure 5.**
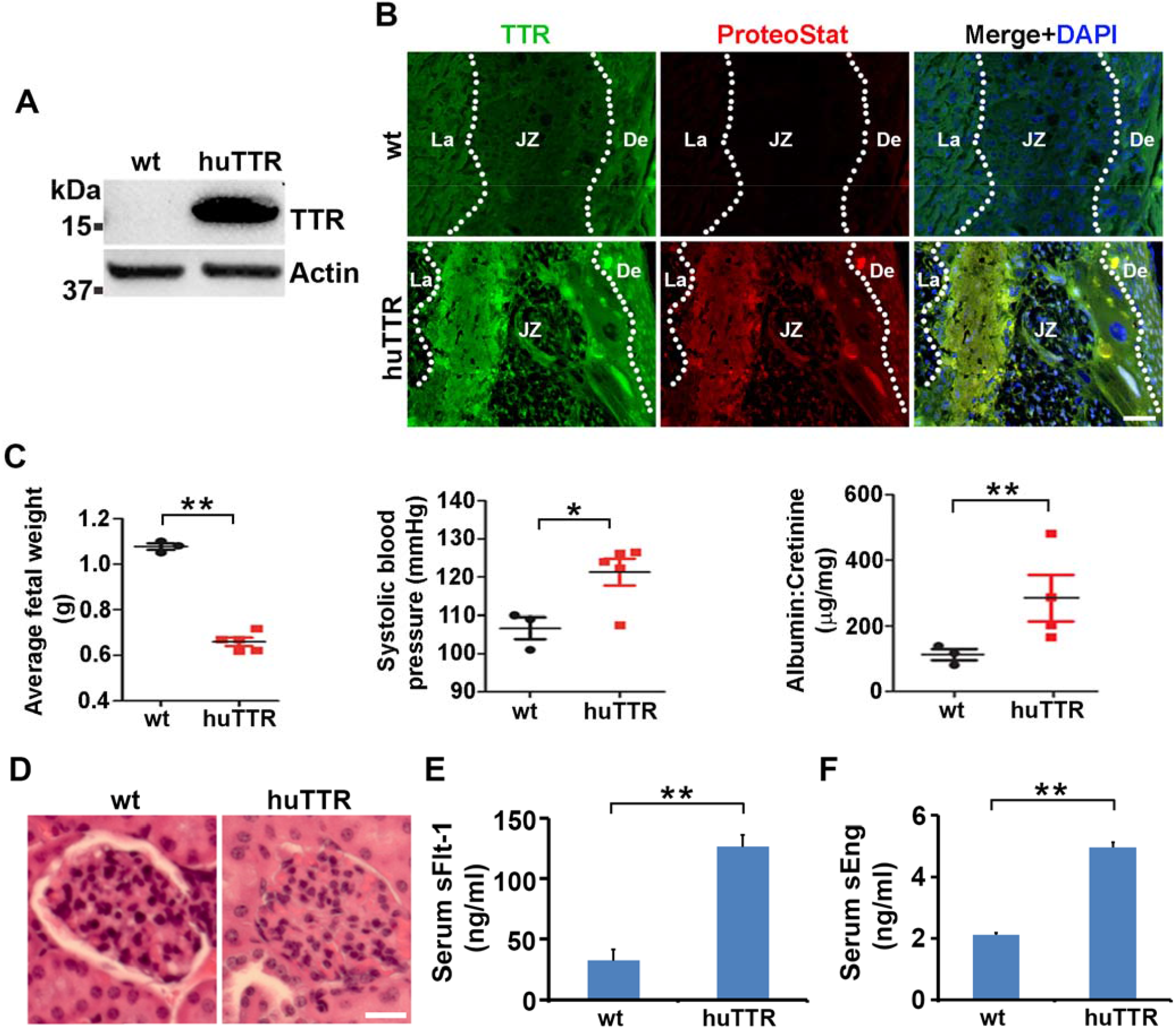
Transgenic mice overexpressing TTR exhibit the deposition of TTR aggregates in the placenta, PE-like features and increased the concentration of serum sFlt-1 and sEng. **A**, Concentration of human TTR in sera from wild-type (wt) and TTR transgenic mice (huTTR). **B**, Colocalization of TTR with ProteoStat fluorescence signal in the placenta of transgenic mice at 7-9 weeks of age. La: labyrinth; JZ: junctional zone; De: decidua. Scale bar: 50 µm. **C**, Comparison of average fetal weight, systolic blood pressure and the ratio of albumin to creatinine between wild-type and transgenic mice. **D**, H&E staining of the kidneys from wild-type and transgenic mice. Scale bar: 60 µm. **E, F**, Concentration of sFlt-1 and sEng in sera from wild-type (wt) and TTR transgenic mice (huTTR). Data were expressed as mean ± SEM and statistically analyzed by one-way ANOVA (n = 3-5). *: *p* < 0.05. **: *p* < 0.01.

Since numerous prior studies have shown that higher serum levels of sFlt-1 and sEng contribute to the pathogenesis of PE^19, 20, 22, 23^, we measured the concentration of these anti-angiogenic factors using respective ELISA kits. The analysis showed significantly higher levels of sFlt-1 and sEng in the sera from pregnant huTTR mice when compared with age-matched pregnant wild-type mice (Figure 5E and F).

To determine if the pregnant huTTR transgenic mice showed alterations in placental perfusion and fetal blood flow, we performed Doppler ultrasound analysis using a Vevo 3100 device (see Methods) and measured peak systolic and diastolic velocity of uterine artery and peak systolic velocity of umbilical artery in pregnant wild type and transgenic mice at gd14 and gd16, respectively. Our results showed decreased uterine and umbilical artery blood flow in huTTR mice compared to wild-type controls (Figure 6).

**Figure 6.**
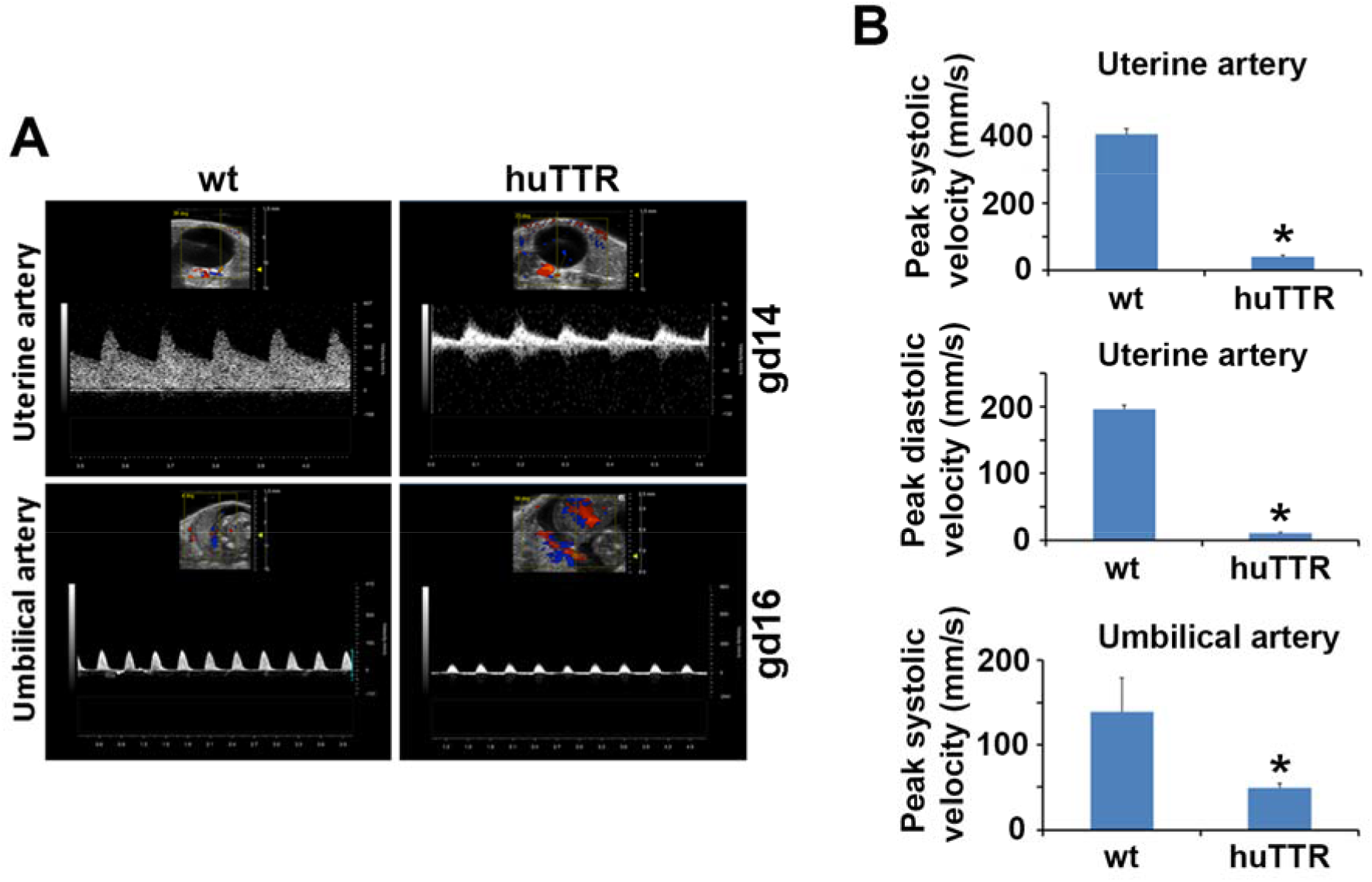
Transgenic mice overexpressing TTR (huTTR) at 7-9 weeks of age display decreased blood flow of uterine and umbilical arteries compared to wild type mice (wt) at gd 14 and gd 16. **A, B**, Doppler ultrasound graph (**A**) and quantitative analysis (**B**) showed the blood flow of uterine and umbilical arteries. Data were expressed as mean ± SEM and statistically analyzed by one-way ANOVA (n = 3). *: *p* < 0.001.

### Loss of mouse TTR function fails to result in PE-like features in TTR knockout mice

The huTTR mice used in Figures 7 and 8 were originally generated on a BALB/DBA background then crossed on to the TTR knockout (129/sv) background over many generations. To evaluate the pregnancy outcomes and the occurrence of PE-like features in *Ttr* knockout mice that did not carry the human *TTR* construct, 7-8-week-old *Ttr* knockout mice were allowed to mate under the same housing and maintenance conditions as the human TTR transgenics. All experimental evaluations were performed as described for the huTTR mice. Our results showed that, consistent with prior studies the TTR knockout mice did not differ from wild type *Ttr* controls with respect to fertility, litter size and fetal weight.^39^ Additionally, we measured blood pressure and serum levels of sFlt-1 and sEng in *Ttr* knockout mice and controls at gd 17. No hypertension was noted. Serum levels of sFlt-1 and sEng were not elevated in the pregnant *Ttr* knockout mice (Figure S6 in the Supplemental Material).

## DISCUSSION

In this study, we observed significant TTR aggregate deposition in placental tissue from women with e-PE. Importantly, we discovered that transgenic mice in which the endogenous *Ttr* genes had been silenced by targeted disruption but overexpressed wild type human TTR, when pregnant experienced PE-like features, exhibited placental deposition of TTR aggregates, and showed high levels of serum sFlt-1 and sEng. Control female pregnant *Ttr* knockout mice that did not carry the normal human gene did not display PE-like features, hence it was unlikely that the PE phenotype was related to the absence of the murine protein. It has been amply demonstrated that despite having similar structure and function, with respect to their capacity to transport thyroxine and retinol binding protein charged with retinol, murine and human TTR differ considerably in thermodynamic and kinetic stability, with the murine tetramer much more stable, showing no dissociation to its constituent monomers under conditions in which the human tetramer readily falls apart. Since it is the dissociated monomer that is prone to aggregation and the formation of toxic oligomers, it is likely that the placental TTR aggregates are a function of the increased amount of human monomer available in these mice and are the cause of the PE-like phenotype.^40^ These changes may explain an observation made early in the generation of the huTTR strain, i.e. that litter sizes from crosses between brother and sister transgenics, in which the surviving pups had very high levels of human TTR, progressively diminished over three generations, requiring maintenance of the transgenic strain in the hemizygous state to preserve the genotype and resulting phenotype.

We also found significant accumulation of TTR aggregates in a cellular model mimicking PE pathology (chronic H/R in PHTs and TCL-1 trophoblasts). TTR aggregates also accumulate in PHTs when they are exposed to drugs inhibiting the proteasome, lysosomal function, or the autophagy pathway. Taken together, these observations strongly suggest that TTR aggregation plays an important role in the pathogenesis of human PE and the role is modulated by placental proteostatic pathways.

It is known that TTR is produced by human trophoblast cells.^28,29^ However, total TTR production is not increased in PE as detected in serum samples from normal pregnancy and PE patients.^11^ Thus, the placental cellular environment in PE becomes one in which the TTR tetramer dissociates and the monomers aggregate.

In the context of these analyses, what factors could be responsible for creating the appropriate environment for protein misfolding and aggregation? Prior studies have shown that hypoxia can initiate ER stress^18^, increase protein misfolding^41^, cause PE-like features in pregnant rats^42^ and IL-10^-/-^ mice^9^, and exacerbate PE symptoms in eNOS^-/-^ mice.^43^ Thus, chronic low oxygen tension or hypoxia, presumably related to diminished placental blood flow, is a key contributing factor to the pathogenesis of PE.^9,18, 42-45^ Our results suggest that the H/R condition seems to contribute to generation of misfolded and aggregated proteins which may in turn induce excessive ER stress and hyper-activated UPR. These conditions most probably lead to the onset of PE-like features.

What is the evidence that TTR aggregation is the result of placental ER stress? During ER stress, BiP is released from its binding site on IRE1α (pIRE1α) to interact with the increased quantity of misfolded proteins, which activates the UPR. However, IRE1α can paradoxically trigger apoptosis by degrading anti-apoptotic microRNAs during excessive ER stress.^46^ Prolonged activation of IRE1α has been shown to induce mutant huntingtin aggregation via the inhibition of autophagic flux, leading to neuronal apoptosis.^47^ PDI, a chaperone of the UPR, maintains -SH group stability, while Ero1 is an oxidoreductase enzyme that catalyzes the formation of protein disulfide bonds in the ER lumen.^48^ Activation of the PDI-Ero1 cycle by misfolded proteins has been shown to lead to the overproduction of reactive oxygen species, oxidative stress and apoptosis.^49,50^ In addition, inhibition of PDI or Ero1 prevents the accumulation of aggregated α-synuclein and promotes cell survival.^50^ These observations suggest that overwhelmed UPR activation is associated with accumulation of protein aggregates and cell death. In our studies, the increased levels of BiP, pIRE1α, PDI, Ero-1, and caspase 3 observed in both PE placental tissue and H/R treated cultured trophoblast cells indicate overwhelmed UPR activation. While BiP is known to bind to protein aggregates, it primarily associates with misfolded polypeptides, serving as a foldase or a holdase. The co-localization of BiP with only a minority of TTR may reflect its differential preference for misfolded over aggregated TTR and the cytoplasmic location of TTR aggregates. The increased amounts of both PDI and Ero-1 in the H/R treated culture trophoblast cells and the PE placenta reinforces the notion of oxido-reductive ER stress, presumably associated with hypoxic state in the pathogenesis of PE. Further, we found that inhibition of IRE1α attenuated Ero1 abundance in the PHTs exposed to H/R. The activation of the PERK pathway is suggested by the increased content of caspase-3 in the H/R treated PHT’s. Taken together, our observations of increased BiP, pIRE1α, PDI, Ero-1 and caspase-3 in the PE placenta and H/R-treated trophoblast cells are consistent with excessive ER stress and hyper-activation of the UPR participating in the pathogenesis of PE. Hyper-activated UPR can inhibit the ability to process misfolded proteins, leading to aggregation of misfolded proteins.

The cytoplasmic TTR aggregates did not co-localize with lysosomal marker, LAMP1. Large protein aggregates are mainly degraded through autophagy-lysosomal machinery under the conditions of mild ER stress.^16^ Our results demonstrated that disruption of the lysosome machinery by chloroquine resulted in accumulation of aggregated TTR in PHT and TCL-1 cells. Thus, we speculate that chronic H/R may impair the autophagy-lysosomal degradation pathway, which inhibits the degradation of aggregated TTR in PHTs. As expected, H/R exposure downregulated the expression of LAMP1, a protein associated with the integrity of the lysosomes, and decreased the co-localization of LAMP1 with TTR aggregates. Lysosomal integrity is essential for the normal functions of autophagic pathways. Therefore, our data suggest that persistent H/R treatment may lead to the accumulation of TTR aggregates by disrupting autophagy-lysosomal pathways.

The e-PE placenta also exhibited higher abundance of SQSTM1/p62, an autophagy receptor that binds ubiquitinated protein aggregates and targets these cargoes to autophagosomes.^51,52^ SQSTM1/p62 is degraded along with ubiquitinated aggregated proteins in the autolysosome after autophagosome-lysosome fusion.^51,52^ Thus, when autophagy-lysosome pathway is disrupted, SQSTM1/p62 accumulates with ubiquitinated protein aggregates. Indeed, we found a larger amount of total ubiquitinated proteins and TTR aggregates in the e-PE placenta vs. control. These findings strongly suggest impairment of autophagy-lysosomal machinery in the placenta from e-PE deliveries, which is consistent with our prior study showing defective lysosome biogenesis and impaired autophagy in severe PE placenta.^16^ Thus, the pathological alterations observed in our cellular model of PE pathology also occur in the e-PE placenta. These molecular events may explain why there is exuberant accumulation of TTR aggregates in the trophoblast layer in the e-PE placenta.

Although TTR aggregation has long been associated with the pathogenesis of hereditary and sporadic forms of systemic amyloidosis, the involvement of TTR aggregation in the pathophysiology of PE has only been recognized recently.^2,11^ High levels of TTR have been reported in amniotic fluid from PE patients and in the placenta from intrauterine growth restriction and severe e-PE deliveries.^53,54^ Our prior studies have demonstrated that sera from PE patients exhibit an increased propensity for TTR-specific protein aggregation, and administration of TTR immunoprecipitated from PE sera induces PE-like features in pregnant IL-10^-/-^ mice.^11^ In the current study, we showed that transgenic mice over-expressing wild type human TTR exhibited robust deposition of TTR aggregates in the placenta, had increased circulating sFlt-1 and sEng, evidence of decreased placental perfusion, and displayed the full spectrum of PE-like features including hypertension, proteinuria and low fetal weight.

Taken together, our findings suggest that TTR aggregation exerts detrimental effects in the placenta contributing to poor trophoblast growth and differentiation. Alternatively, it can be speculated that rather than solely being a consequence of placental hypoxia, toxic protein aggregates dysregulate placentation, trophoblast invasion and spiral artery remodeling and increase the production of pregnancy-compromising substances, such as anti-angiogenic factors, extracellular vesicles and alarmins, which may lead to endothelial dysfunction and systemic inflammation as the precise contributory mechanisms of PE.

## PERSPECTIVES

We and others recently demonstrated that toxic protein aggregation (proteinopathy) is associated with the pathogenesis of PE. However, it is not clear what altered pathways in the placental microenvironment may lead to protein aggregation and what proteins may be the target of such activity. It is well known that human TTR is synthesized by trophoblast cells. It is also clear that the human TTR tetramer can dissociate to release its aggregation prone monomer. We have shown that various forms of ER stress result in TTR aggregation in trophoblast derived cells. We have also shown that pregnant mice transgenic for multiple copies of the wild type human TTR gene with all the elements required for tissue specific expression and having high levels of human TTR, develop placental pathology, serum markers and abnormalities of placental blood flow that are typical of human PE, with no other manipulation other than the stress of pregnancy. This study demonstrates for the first time that accumulation of TTR aggregates in the e-PE placenta may be caused by excessive ER stress, hyper-activated UPR and impaired autophagy-lysosomal machinery. Moreover, our results suggest that TTR aggregates may, at least in part, be implicated in the onset of PE-like features. Our studies further show that TTR aggregation is prevalent in the junctional zone of the placenta of huTTR mice and causes elevated production of sFlt-1 and sEng. In our studies, loss of mouse TTR function in TTR knockout mice did not impact fertility and normal pregnancy outcome, rather it was the gain of toxic function of the TTR aggregates responsible for the PE-like phenotype. The orally administered TTR stabilizer tafamidis has been shown to be both safe and effective for the treatment of the TTR amyloidoses. It will be interesting to see if administration of the compound to the human TTR transgenic mice will modulate or prevent the PE phenotype. Alternatively, targeting protein aggregation and restoration of autophagy could offer other potent pharmacological options for PE.

## Novelty and Significance

### What Is New

- We demonstrate that TTR aggregates accumulate in the placenta from PE patients and huTTR mice.
- Accumulation of TTR aggregates can be recapitulated in primary human trophoblasts treated with ER stress inducers, autophagy-lysosomal disruptor or chronic H/R.
- Pregnant transgenic mice overexpressing wild-type human TTR expressed in the human tissue specific manner demonstrate robust accumulation of TTR aggregates in the placenta, show significantly higher levels of serum sFlt-1 and sEng, and display a PE-like clinical phenotype.
- Loss of mouse endogenous TTR function fails to result in the onset of PE-like features.

### What Is Relevant

- Excessive ER stress, overwhelmed UPR, and impaired autophagy-lysosomal pathways can lead to the accumulation of TTR aggregates in the PE placenta.
- TTR aggregates play a causative role in the pathogenesis of PE.

### Summary

Chronic low oxygen tension may induce excessive ER stress, hyper-activate the UPR and impair the autophagy-lysosomal pathways, leading to the accumulation of TTR aggregates in PHTs and PE placenta. Toxic protein aggregates may, at least in part, contribute to the pathophysiology of PE. It can be speculated that controlling ER stress and excessive UPR and restoring autophagy-lysosomal pathways in the placenta can inhibit protein aggregation. These pathways can be targeted to have new therapeutic avenues for preventing and treating this severe pregnancy complication.

## Data Availability

All data produced in the present work are included in the manuscript.

## Nonstandard Abbreviations and Acronyms

ER: Endoplasmic reticulum
BiP: Binding immunoglobulin protein
Ero1: endoplasmic reticulum oxidoreductin 1
IRE1α: Inositol-requiring enzyme-1α
LAMP1: lysosome-associated membrane protein 1
PDI: protein disulfide isomerase
pIRE1α: Phosphorylated inositol-requiring enzyme-1α
sEng: soluble endoglin
sFlt-1: soluble fms-like tyrosine kinase-1
TTR: transthyretin
UPR: unfolded protein response

## Acknowledgments

We thank the Department of Pediatrics, Women & Infants’ Hospital of Rhode Island, Warren Alpert Medical School of Brown University, and Center of Biomedical Research Excellence (COBRE) Core Facility for the continued support.

