## Supplementary Material for "Evidence from human placenta, ER-stressed trophoblasts and transgenic mice links transthyretin proteinopathy to preeclampsia"

### Supplemental Material

### Supplemental Figures and Figure legends

#### Figure S1

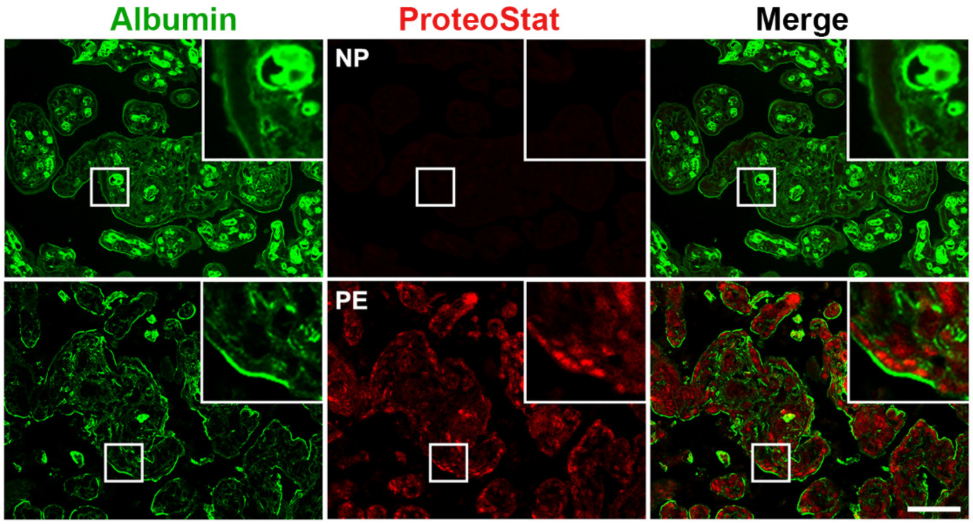


**Figure S1. Albumin does not co-localize with ProteoStat fluorescence in the placenta from e-PE and control**. Placenta sections were immunostained with anti-albumin antibody (green) and co-stained with ProteoStat dye (red). Nuclei were stained with DAPI (blue). The images were representatives of at least 3 independent experiments. Scale bar: 20 μm

#### Figure S2

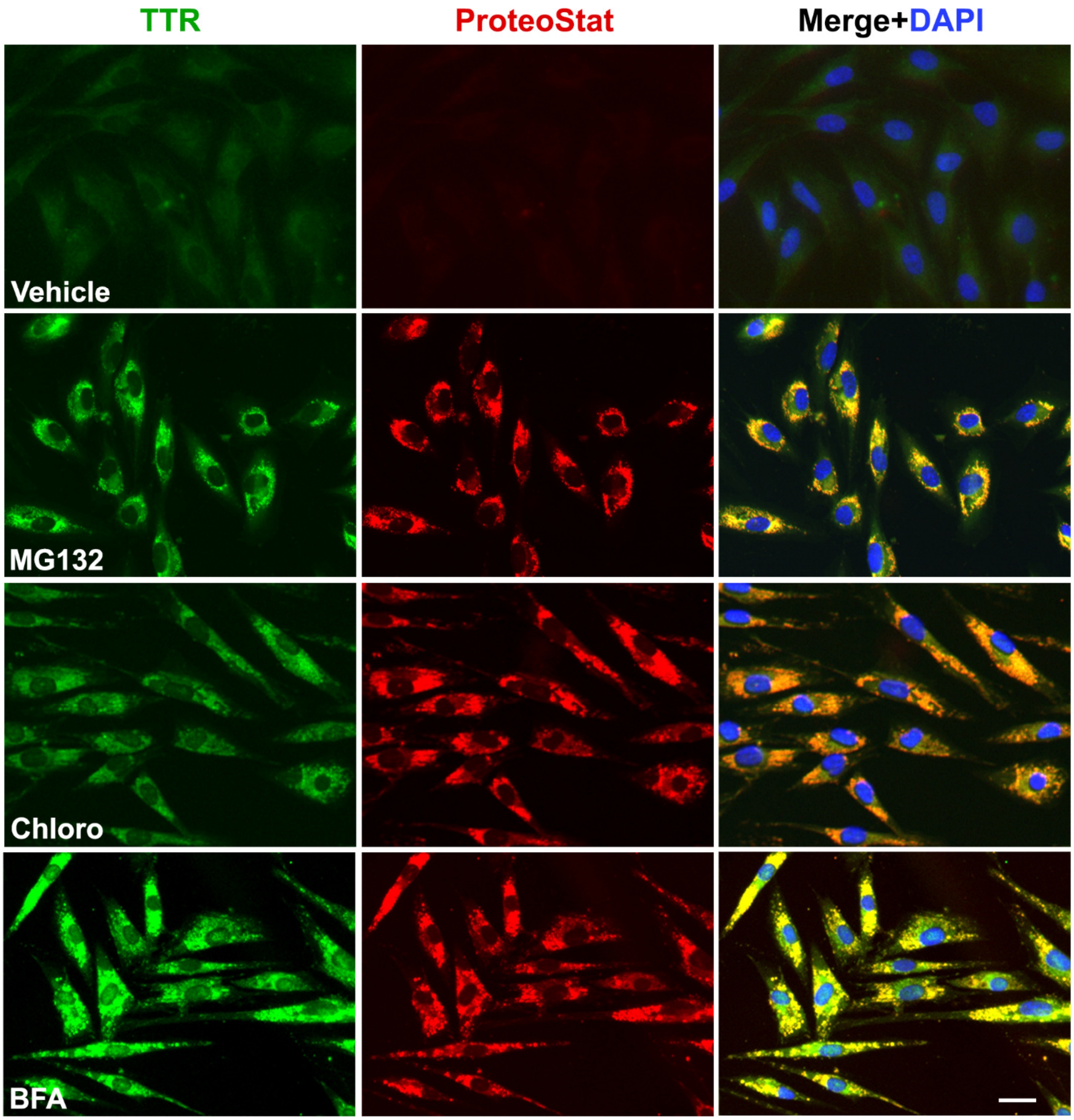


**Figure S2. MG132, Chloroquine and BFA induce the accumulation of TTR aggregates in PHTs.** The cells were treated overnight with MG132 (10 µM), chloroquine, (10 µM) or BFA (2 µg/ml), fixed and then immunostained for TTR (green) and co-stained with ProteoStat dye (red). Nuclei were stained with DAPI (blue). Colocalization of TTR with ProteoStat dye showed yellow in color after these two channels were merged. The images were representatives of at least 3 independent experiments. Scale bar: 20 μm

#### Figure S3

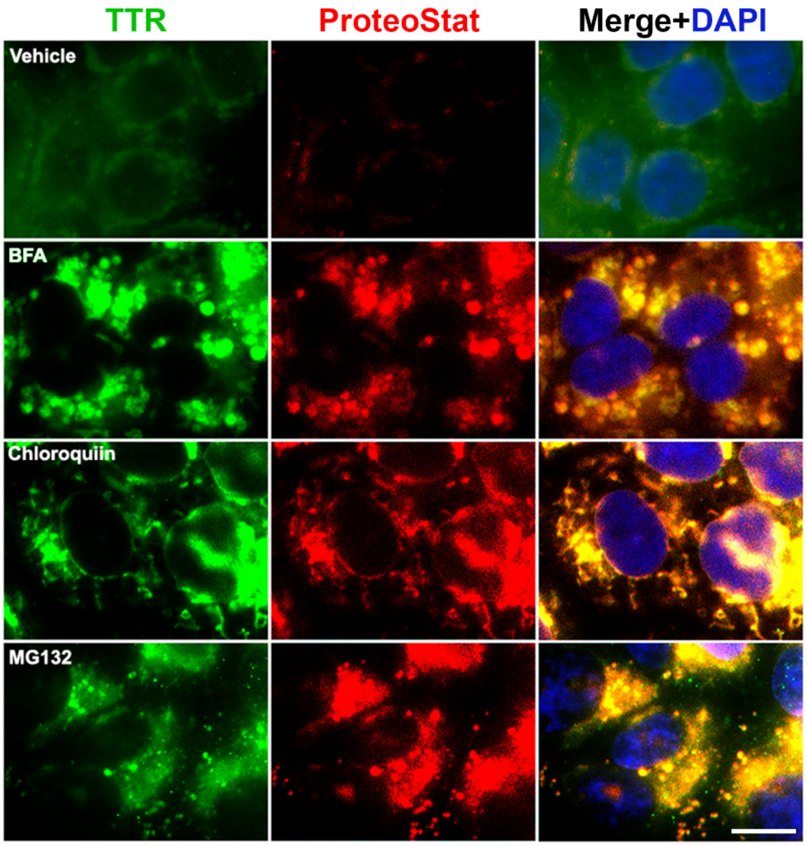


**Figure S3. MG132, Chloroquine and BFA induce the accumulation of TTR aggregates in TCL-1 trophoblasts**. The cells were treated overnight with MG132 (10 µM), chloroquine, (10 µM) or BFA (2 µg/ml), fixed and then immunostained for TTR (green) and co-stained with ProteoStat dye (red). Nuclei were stained with DAPI (blue). Colocalization of TTR with ProteoStat dye showed yellow in color after these two channels were merged. The images were representatives of at least 3 independent experiments. Scale bar: 20 µm

### Figure S4

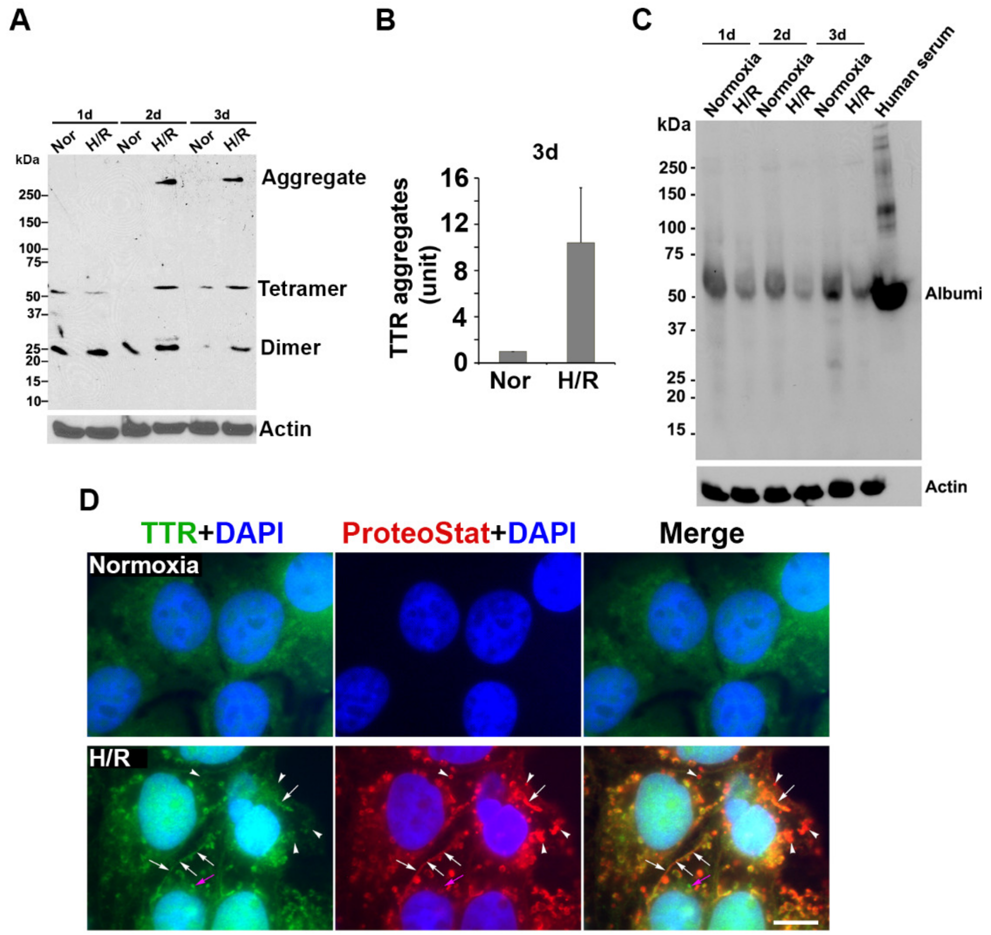


**Figure S4. H/R induces the accumulation of TTR aggregates in TCL-1 trophoblasts**. The cells were treated with normoxia or H/R and harvested at various time points. **A, B,** Western blotting under non-reducing conditions and quantitative analysis showed higher levels of TTR aggregates in H/R-treated TCL-1 cells ($p < 0.01$, n = 3). **C**, Western blotting under non-reducing demonstrated that H/R did not induce albumin aggregation. Human serum was used as a positive control. **D,** Representative images showed the co-localization of robust TTR and ProteoStat signals in H/R-exposed TCL-1 cells. The nuclei were stained with DAPI (blue). Scale bar: 20 μm

### Figure S5

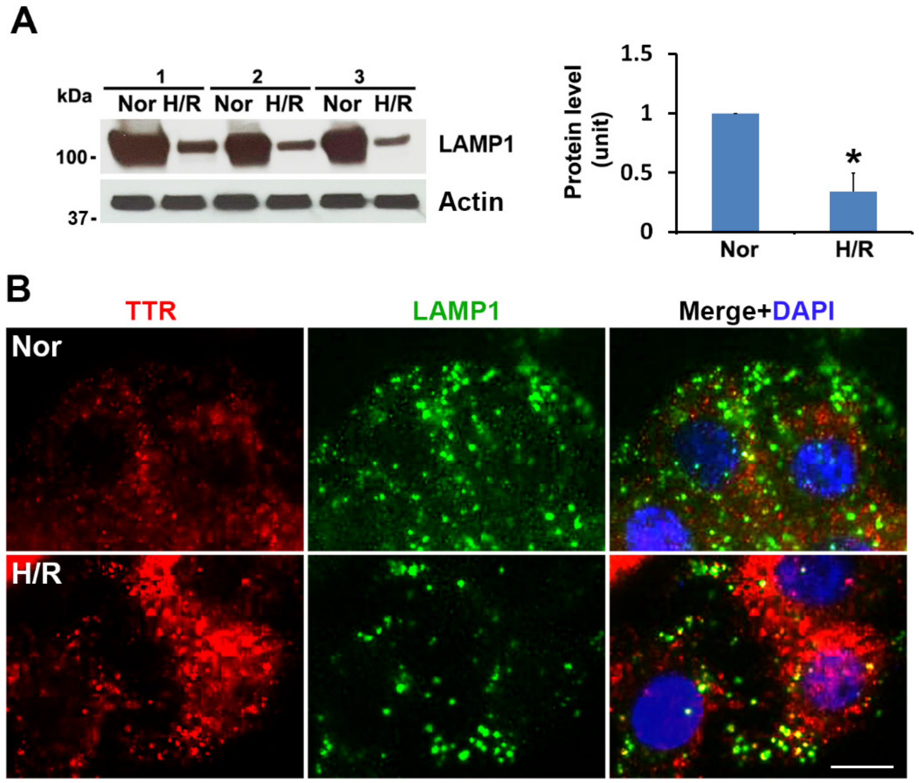


**Figure S5**. **Effect of H/R treatment on the expression of LAMP1 and its colocalization with TTR aggregates in PHTs.** The cells were treated with normoxia or H/R for 3 days, lysed for western blotting or fixed for immunostained for TTR (red) and LAMP1 (green). **A,** Western blotting of LAMP1 and quantitative analysis. Data were expressed as mean ± SEM and statistically analyzed by the Student's *t*-test (n = 3). *: $p < 0.05$. **B,** Colocalization of TTR with LAMP1. The nuclei were visualized with DAPI staining. The images were representatives of at least 3 independent experiments. Scale bar: 20 µm

### Figure S6

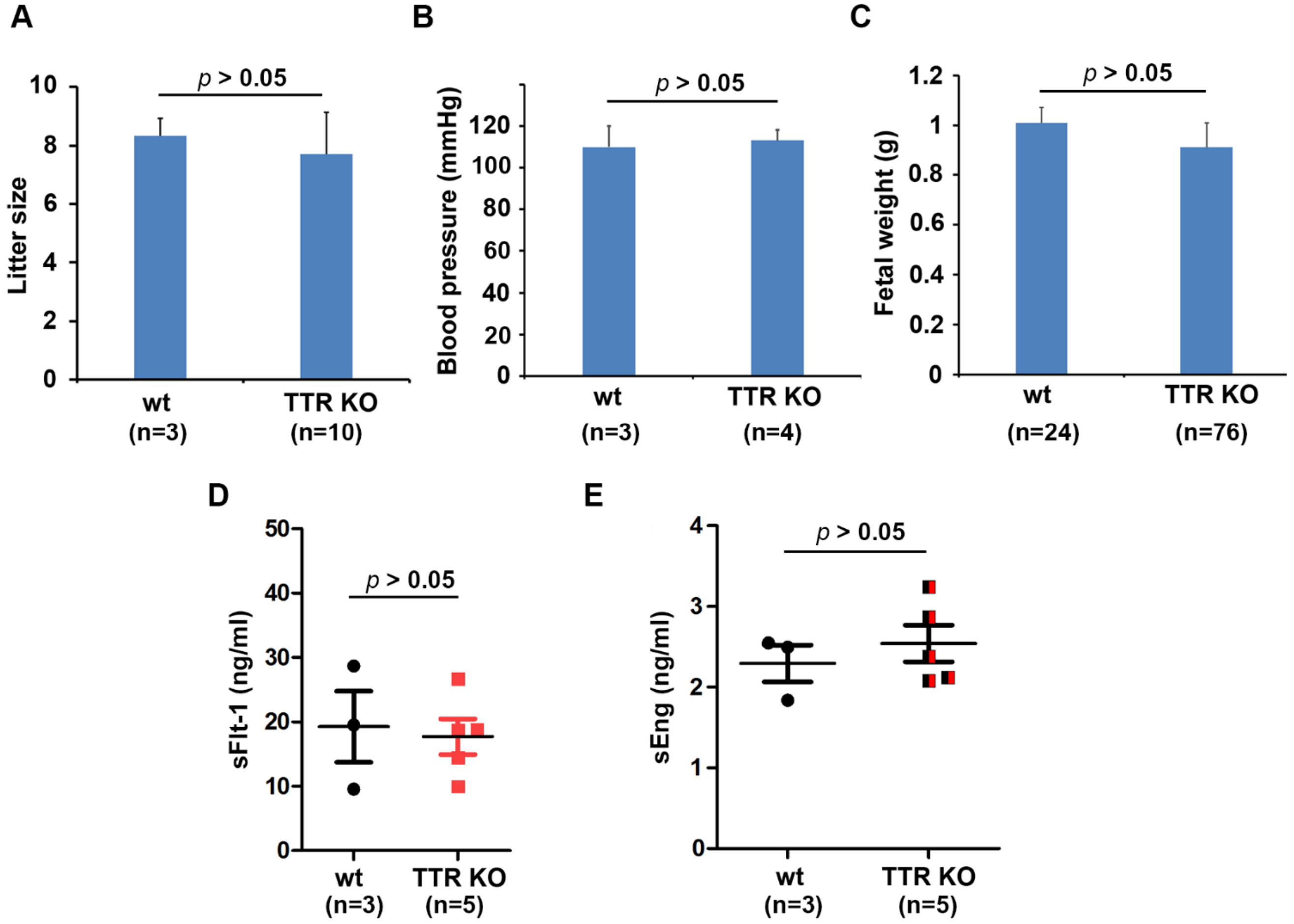


**Figure S6. Knockout of mouse endogenous TTR fails to induce PE-like features in pregnant mice at gd 17.5.** Mice with TTR knockout (TTR KO) did not exhibit differences in litter size (**A**), blood pressure (**B**), fetal weight (**C**), and serum levels of sFlt-1 (**D**) and sEng (**E**) compared to wild-type controls (wt). $p > 0.05$

**Supplemental Table**

**Table S1: Demographic and clinical characteristics of patients**

| Variable | Early onset preeclampsia (n=8) | Gestational age-matched control (n=8) | *p*-value |
|---|---|---|---|
| Age (years) | 29.8 (7.1) | 24.8 (5.8) | † 0.41[1]<br>†† 0.14[1] |
| Race<br>White<br>Black<br>Hispanic<br>Other | <br>4 (50%)<br>1 (12.5%)<br>3 (37.5%)<br>0 | <br>3 (37.5%)<br>2 (25%)<br>3 (37.5%)<br>0 | <br>†0.51[2]<br>†† 0.84[2] |
| BMI (kg/$m^2$) | 32.0 (6.2) | 31.5 (7.2) | † 0.10[1]<br>†† 0.87[1] |
| Gestational age at delivery (weeks) | 31.7 (1.5) | 33.1 (1.3) | † 0.73[3]<br>†† 0.06[3] |
| Maternal temperature (°C) | 36.9 (0.13) | 36.8 (0.18) | † 0.23[1]<br>†† 0.36[1] |
| Maximum systolic blood pressure (mmHg) | 179 (13.2) | 122 (13.8) | † < 0.001[3]<br>††<0.001[3] |
| Maximum diastolic blood pressure (mmHg) | 113 (5.9) | 74.6 (5.8) | † < 0.001[3]<br>††<0.001[3] |
| Mode of delivery<br>Vaginal<br>Cesarean Section | <br>3 (37.5%)<br>5 (62.5%) | <br>5 (62.5%)<br>3 (37.5%) | <br>† 0.5[2]<br>†† 0.62[2] |
| Maternal hemoglobin (g/dl) | 10.4 (0.9) | 10.5 (1.6) | † 0.32[1]<br>†† 0.81[1] |
| Maternal platelets (x$10^3$/µl) | 101 (68.3) | 195 (71.2) | † 0.05 [1]<br>†† 0.02[1] |
| AST (U/L) | 247 (288) | 11 – 30* | N/A |
| Serum creatinine (mg/dl) | 0.97 (0.25) | 0.5 – 1.1* | N/A |
| Urine protein : creatinine | 5.0 (6.6) | < 0.3* | N/A |
| Birth weight (grams) | 1334 (129) | 2133 (382) | † 0.11[1]<br>††<0.001[1] |

Data presented as Mean (standard deviation) for continuous variables
Data presented as n (%) for categorical variables
*AST, serum creatinine, and urine protein:creatinine not measured in Control subjects and presented as normal ranges.
[1]t-test
[2] Fisher's exact test
[3] Wilcoxon rank-sum
†late onset preeclampsia with severe features versus term controls

†† early onset preeclampsia with severe features versus preterm controls
